# IDoser: Improving individualized dosing policies with clinical practice and machine learning

**DOI:** 10.1101/2023.03.28.23287859

**Authors:** Nuria Correa, Jesus Cerquides, Rita Vassena, Mina Popovic, Josep Lluis Arcos

## Abstract

**Background:** Finding the correct drug dose for a specific condition is a key step in many treatments, and failing to do so can lead to deleterious consequences to patient health. Clinical protocols are derived from drug development phase prospective trials. While carefully designed, these often do not include all potential patients, comorbidities or clinical outcomes, ultimately leading to sub-optimal dosing policies. Observational datasets provide real-world information that cannot be substituted with data collected in a controlled environment. Several published methodologies have applied observational datasets for the development of clinical protocols, however these are only applicable whenever these datasets are varied and complete. Often, clinical observational datasets do not comply with these requirements. Computational methods can and should exploit field knowledge to address weaknesses associated with clinical observational data.

**Methods:** This paper proposes IDoser, a core dosing model that links drug dose to relevant covariates via a set of coefficients, and includes a loss function to codify needed assumptions and requirements. Coordinate descent is used to obtain a fitted model with minimal loss. The loss function is also used to measure performance when validating the model with unseen data. Our proposal is validated using the case of follicle stimulating hormone (FSH) dosing for controlled ovarian stimulation (COS).

**Results:** The proposed Individualized Doser (IDoser) achieved significant improvements when loss values were compared to observed clinical practice and a selected literature benchmark and during the validation phase.

**Conclusions:** This methodology constitutes a simple but effective method to bridge the gap between current clinical dosing policies and gold policies based on the true underlying and often unknown dose-response functions.

## 1. Introduction

Once a patient decides to put their health in the hands of a clinical professional for a needed and/or wanted treatment, they rightfully expect to receive the best possible care. One of many key steps in a given treatment includes the selection of the right dose for a prescribed drug. This dose will be determined with an optimal outcome in mind, for example that the patient’s blood pressure is balanced, or that their temperature lies between certain values. All available knowledge on the underlying dose-response relationship will be used by clinical professionals to determine the right dose of drugs to achieve optimal outcomes. However, this relationship is often not wellknown, and may result in suboptimal dose selection, ultimately leading to subpar clinical care for the patient.

An example of this situation is the empirical case covered in this research, which pertains to finding the right first dose of follicle stimulating hormone (FSH) for controlled ovarian stimulation (COS). COS is a key step in the treatment of infertility, used in women to induce the ovary to develop multiple follicles and eggs (oocytes) simultaneously. Once the follicles reach the appropriate size, they are punctured and aspirated in a simple surgical procedure and ultimately mature oocytes are collected. An appropriate COS is critical to the success of in vitro fertilization (IVF), as the number of mature oocytes collected is tightly associated with the chances of achieving a pregnancy safely. As with many clinical procedures, there is an optimal outcome (10 to 15 mature oocytes [34, 40]). Lower or higher outcomes are undesirable, as a lower number of mature oocytes is associated with lower chances of achieving a pregnancy, while a higher number of mature oocytes increases the risk of ovarian hyperstimulation syndrome (OHSS), a serious adverse complication resulting from an exaggerated response to excess hormones.

Individuals with similar characteristics (sometimes even the same individual at different points of time) can respond differently to the same treatments – a direct consequence of the lack of knowledge surrounding the dose-response relationship, as well as its interaction with unknown factors. Such cases lead to suboptimal outcomes, as experienced in FSH dose prescription.

Pharmacokinetic/pharmacodynamic (PK/PD) studies constitute an important step in drug development and clinical approval. These studies inform the design and execution of Randomized Controlled Trials (RCTs), in which the drug is tested in clinical conditions. Clinical protocols are then derived from the results of RCTs to guide practice. Ultimately, results range from acceptable to almost optimal, depending on the fit of the PK/PD models on the final objective population and real clinical outcome. The final unexplained variability constitutes the aim of further research. This variability is mainly related to the real target population distribution and its accompanying comorbidities.

While this system allows for most patients to receive an adequate dose of drug, dose protocols or policies may not be fitting all patients evenly. Clinicians use their own experience and published literature to fill in the knowledge gaps, however this is still not optimal. There are ways to adapt these initial models to real-world clinical use, but either imply using new prospective data or very diverse observational datasets. The first option involves prospectively testing different dose concentrations on the same individual. The second option remains difficult to define. While clinicians do adapt protocols when necessary, they seldom deviate from them, generating data with sparse diversity.

In this research we aim to present a straightforward methodology to improve individualized drug dosing policies using available observational datasets and field knowledge, while simultaneously incorporating requirements of the specific problem.

## 2. Related work

PK focuses on the relationship between the drug dose administered and its concentration in different body compartments at specific time points. Conversely, PD focuses on what the drug does to the body, namely in the exposure to effect relationship [28]. Model-Informed Precision Dosing (MIPD)[24, 9] constitutes a good approach to individualize dose protocols, and its ideal form passes through obtaining well fitted models for both PK and PD of the studied drug. Drugs approved for clinical use habitually have a published PK/PD model, derived from phase III clinical trials.

Nevertheless, these models are not applicable to all patients; first, phase III PK/PD studies tend to exclude biomarkers known to affect the drug, although it is relatively common for individuals to have multiple comorbidities [14]. Second, only a relatively selected patient population tends to be included in these trials. Third, uncritically applying these models assumes that the target population parameters have the same distributions as the study sample, often an incorrect assumption due to, for instance, socioeconomic status, genetic and ethnic dispersion and geographical distribution, amongst others [24]. Finally, it is not uncommon to find PK/PD models fitted for outcomes that are different from the objective of clinical interest, or unrelated to key biomarkers known to affect the individual dose-response function variability. This is the case for FSH dose in COS [35, 3, 1].

When the developed model is not applicable to the general population, the use of non-linear mixed methods [38, 39], physiologically based PK models (PBPK) [21], Bayesian methods [37, 9, 15], and traditional PK/PD methods combined with machine learning (ML) [32] can be applied to ameliorate the prediction. However, to be improved, all of these approaches require an available covariate linked PK/PD model to be improved as a starting point. Alternatively, a PK/PD model can be developed, yet this approach is both computationally and labor-intensive, while often requiring data on drug blood concentrations after treatment, which may not be available [31, 25]. These requirements are sometimes not met in clinical practice. Nevertheless, the reliance of these methods on physiological and pharmacological concepts is a great advantage for the explainability and trustworthiness of the resulting models.

Alternatives are available for single dose observational data, including the combination of ML and causal inference methodologies, as presented by Bica et al. [5]. For binary treatments, the propensity score (probability of an individual of receiving certain treatment) is used to adjust for treatment selection bias. For multiple or continuous treatments this concept is translated to the generalized propensity score (GPS)[19, 16]. This score is used to weight samples while estimating the outcome value. Unfortunately, propensity score models must be very well determined and can be numerically unstable due to extreme propensity weights [4]. Recent methods to ameliorate this problem include kernel functions to estimate the GPS [8, 22] and Doubly Robust (DR) ML to estimate outcome values. Other works discretize the treatment space [6, 36], or use generative adversarial methods [5]. These are very good approaches for estimating dose-response relationships but require making two assumptions: positivity or overlap (every individual has non-zero probability of receiving every treatment option) and unconfoundedness (all treatments and outcomes affecting variables are accounted for). In a clinical settings, these assumptions can be very challenging to comply with, and certainly cannot be assumed in the case of FSH use.

Two nomograms [26, 11], and a model based on a multivariate regression [17] have been published, in an attempt to optimize individualized FSH dosing policies without using PK/PD models. All three studies used known relevant biomarkers: age of the patient, anti-Müllerian hormone (AMH), antral follicle count (AFC) and basal FSH levels. Subsequent RCTs to test their efficiency showed a reduction in Ovarian Hyperstimulation Syndrome (OHSS) on one end [33], and an increased proportion of patients with an optimal range of outcome on the other [2], while the mean number of oocytes was not significantly different. Unfortunately, these models were developed only for normo-ovulatory women under the age of 40 years, effectively leaving out the most difficult patients to dose.

## 3. The Individualized Dose Improvement Problem

To formalize the problem under study, next we introduce the Individualized Dosage Improvement Problem (IDIP). Given a large population of *N* patients *P*, the goal of an IDIP is to select the optimal quantity of a certain drug, which we refer to as the *dose*. For every patient *p*_*i*_ ∈ *P* a *dose* and its outcome or *response* is recorded, and there is only a pair of *dose-response* values. We represent the response of *p*_*i*_ by a real number which we refer to as *y*_*i*_ ∈ ℝ, and the dose as *d*_*i*_∈ [0, ∞). We assume that the response can be measured by a single real number. Furthermore, we assume that the desired levels of response are known for each individual, which we describe as 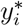.

Each patient *p*_*i*_ ∈ *P* is described by a set of *k* characteristics 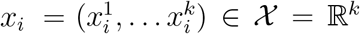. These characteristics can include for example, the patient age in years, its weight, height, gender, values of previous analysis, and so on.

An *individualized dosage policy*, or IDP, *π* : X → ℝ is a function that decides a *proposed dose* 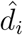 provided the characteristics of the patient (*x*_*i*_).

The objective is to find an IDP or *π* with the minimum error or *loss* possible. Mathematically, this means identifying

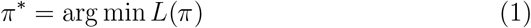

where *L* is a *collective loss* function. That is, *L*(*π*) represents the quality of an IDP *π*. The collective loss is computed as a sum of *individual losses* (*l*_*i*_), one for each patient *p*_*i*_. The individual loss of a dose on a patient 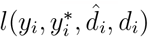 measures how good a proposed dose 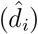 would be depending on the real *d*_*i*_, its correspondent *y*_*i*_ obtained and the objective 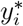. Thus, the *Collective loss* can be mathematically defined as

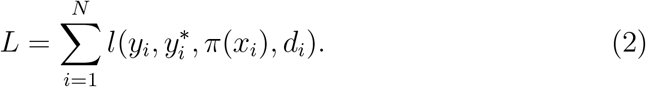

In the IDIP, we are provided with data that describes current practice for the dose policy of the drug. Specifically, we are provided with information about *N* patients out of the complete population, and for each patient *p*_*i*_, *i* ∈ [1..*N*], that has been administered the drug, we record

- their characteristics *x*_*i*_ ∈ X ;
- the dose of drug administered to this patient, namely *d*_*i*_ ∈ [0, ∞);
- the response value obtained, namely *y*_*i*_ ∈ R.

The main challenge posed is: How can we use the information available from current dosing practices to create an individualized dose policy with minimal loss?

## 4. Proposal: Individualized Doser (IDoser)

Our proposal hinges on the following two assumptions:

1. The dose-response function is *monotonic*, that is the larger the dose, the bigger (or equal) the expected response.
2. There is a known *optimal outcome* (*y*^*\**^) and it is known to us. This can either be a range or a point.

*IDoser* is then constructed around two elements: (1) A **Core dosing model** that relates *X* to *d* through a set of coefficients that we describe as *γ*, and is used to predict 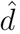; and (2) A **loss function** that evaluates the predicted 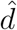 depending on *d* and *y* (see Figure 1).

**Figure 1:**
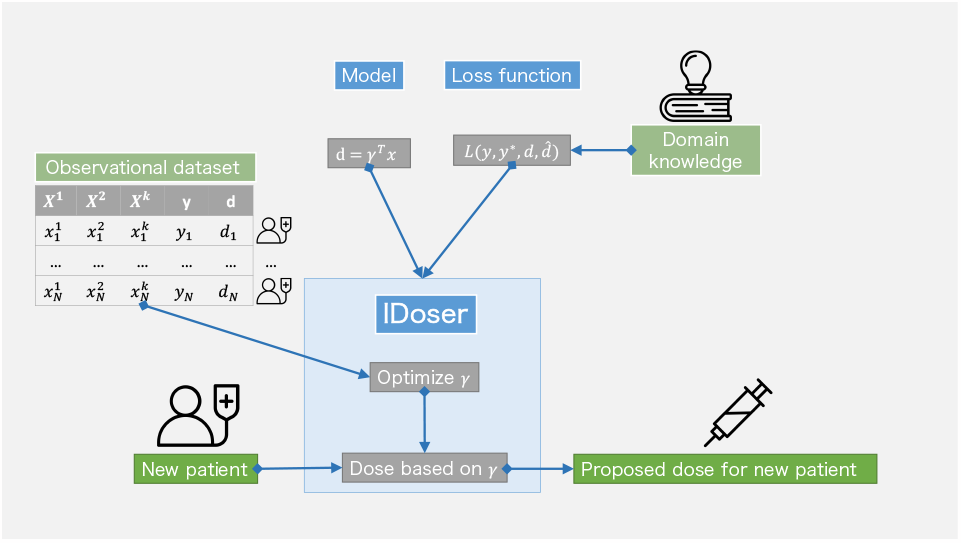
Principal components of IDoser.

We will review both elements in the following subsections. For the rest of the manuscript positive monotonicity will be the default assumption. Nevertheless, IDoser can be easily adapted for a negative monotonicity assumption.

### 4.1. The core model

Given that our main interest lies in predicting the optimal dose for each patient, a general and basic core model is represented as follows:

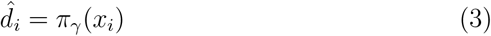

And can be specified in its simplest lineal form as:

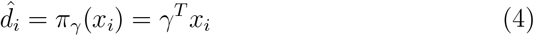

This is the simplest form that complies with the monotonicity assumption and the requirement of relating the dose 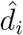 to the covariates *x*_*i*_ through *γ*, but other more complex forms can be used as needed.

### 4.2. Loss function

Being able to evaluate a hypothetical or counterfactual dose is key for achieving an improvement on any real dosing policy (*π*). For this purpose, we codify evidence-based knowledge into the loss function. Namely, our two assumptions: positive monotonicity, and the existence of a desired outcome *y*^*\**^. This can be easily introduced as follows:

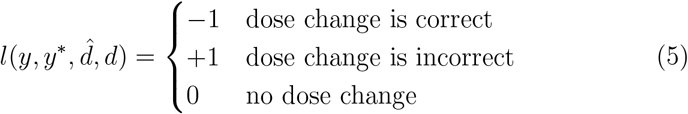

where a correct dose change is increasing *d* whenever *y < y*^*\**^, and decreasing *d* when *y > y*^*\**^. Any change outside of these assumptions would be incorrect (Figure 2).

**Figure 2:**
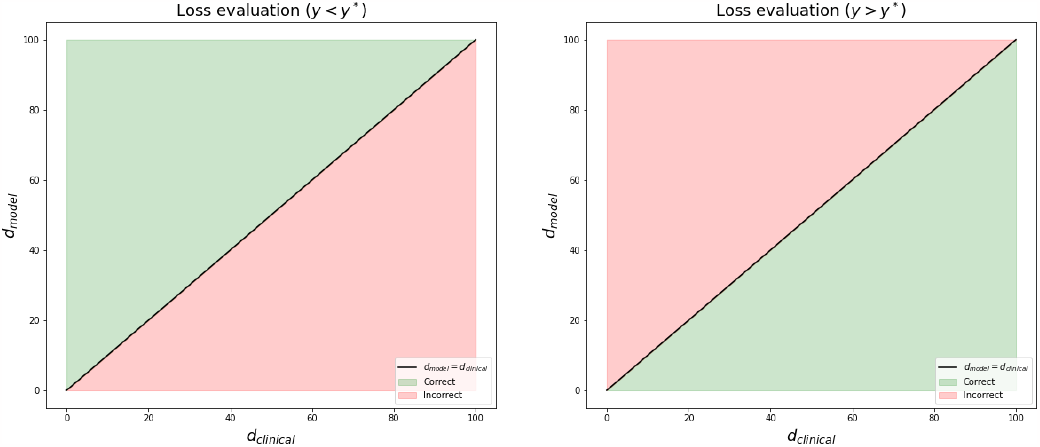
Graphical representation of loss evaluation for cases where *y < y*^*\**^ (left) and where *y > y*^*\**^ (right)

Given positive monotonicity, for any *p*_*i*_ that has *y*_*i*_ *> y*^*\**^, an increase on dose would move *y*_*i*_ further from *y*^*\**^, hence impairing the outcome. In the situation, an improvement would be to reduce dose. On the situation where *y*_*i*_ *< y*^*\**^ the reverse is true.

The function that generates the loss evaluation can be modified as needed for negative monotonicity and, further, complemented with additional rules that may be required depending on the situation or specific use cases.

One example of this is not considering any change in the right direction as good, and introducing limitations on dose change. This kind of limitations would ensure that uncertainty is considered, as larger changes in dose imply less confidence in its effect.

Another rule may involve introducing a certain threshold to start considering 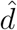 different from *d*. These additional rules would consequently change then our allocation of loss value as represented in Figure 3 and will be exploited in our use case.

**Figure 3:**
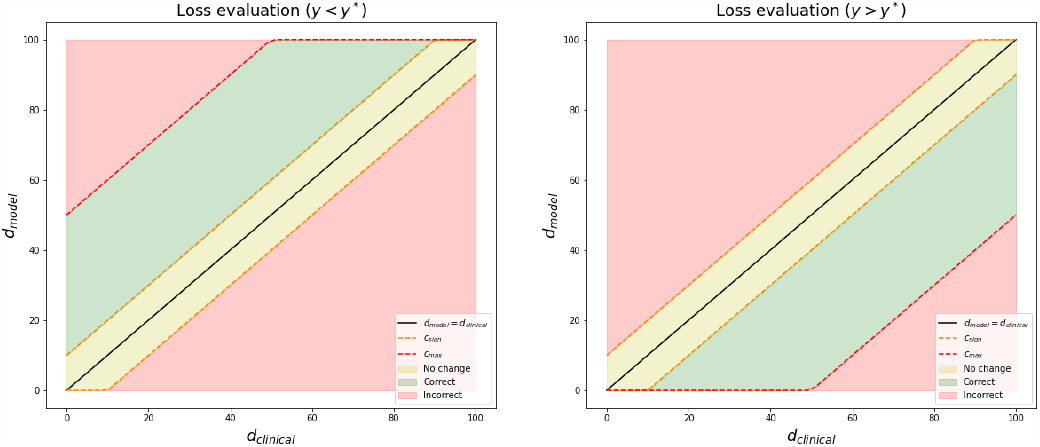
Graphical representation of loss evaluation for cases where *y < y*^*\**^ (left) and where *y > y*^*\**^ (right) with additional rules considering maximum change allowed and a minimum change threshold.

### 4.3. Optimization of parameters

After definition of the core model and the loss function according to the use case selected, the parameters of the model are found by minimization of the loss function. Here, we propose a coordinate descent algorithm [44], to iteratively establish the set of parameters that results in a minimum collective loss or *L*:

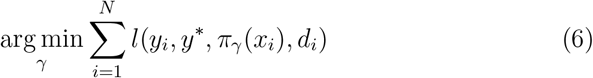

Once a minimum is reached within a randomly selected portion of the database (training), the resulting *γ* parameters and the core model associated with them are considered to be the optimized dosing policy *π*^*\**^.

## 5. Use case

The use case aims to find the right FSH dose in a COS for an IVF treatment.

In the available observational dataset, a set of covariates related to the ovarian reserve of the patient are observed, together with the dose of FSH prescribed by clinicians and the outcome, measured in the number of mature oocytes retrieved. The covariates include: patient age at the time of treatment, body mass index (BMI), AFC, AMH levels, and basal endogenous FSH levels. Two databases were retrieved. One dedicated to developing the dosing models, composed of first IVF cycle patients undergoing treatment between January 2011 and December 2019; and a second one reserved only for validation of the resulting models (cases from January 2020 to September 2021). A summary of the characteristics of the two databases can be found in Table 1.

**Table 1:** Summary statistics of development and validation databases.

|  | Development database<br>(n = 7768) |  | Validation database<br>(n = 273) |  |
| --- | --- | --- | --- | --- |
| **age** | $37.09 \pm 4.85$ | [18-51] | $38.13 \pm 4.10$ | [24-46] |
| **BMI** | $23.75 \pm 4.22$ | [14.53-45.18] | $22.98 \pm 4.02$ | [16.45-41] |
| **AFC** | $11.92 \pm .7.73$ | [0-81] | $11.49 \pm 9.15$ | [0-85] |
| **AMH** | $2.38 \pm 2.33$ | [0.01-32.95] | $2.29 \pm 2.5$ | [0.01-23.70] |
| **basal FSH** | $7.47 \pm 4.19$ | [0.1-94.00] | $8.78 \pm 6.72$ | [0.93-89.60] |
| **FSH dose** | $246.96 \pm 58.95$ | [100-600] | $268.64 \pm 54.73$ | [112.5-450] |
| **MII** | $7.30 \pm 5.26$ | [0-47] | $6.55 \pm 6.07$ | [0-36] |

Thanks to available literature, we confirmed that both assumptions needed for our proposal hold true. For assumption 1 (positive monotonicity), while some evidences in cows may defy it [23], in the human species, no increment of FSH dose results in a lower number of oocytes retrieved under the same circumstances (same patient, same menstrual cycle) [35, 3, 27, 1]. The only negative effects of higher doses of FSH observed in human relate to the quality of oocytes [29] and not their quantity. As such, the positive monotonicity assumption holds. This is not to say that oocyte quality should be disregarded, rather that both quality and quantity are relevant for the cycle success, given that only collected and fertilized oocytes have the chance, by definition, to develop into a blastocyst stage embryo [30, 43]. Therefore an equilibrium must be sought by defining an optimal number of oocytes to be achieved. For assumption 2 (known optimal outcome), clinicians select the first dose of FSH in order to obtain an optimal number of mature oocytes that is known for all patients, although there is some discussion around what constitutes an optimal number in literature [41, 34, 40, 20, 7]. In this paper, we have defined it to be between 10 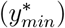 and 15 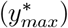 mature oocytes, following the recommendations by Sunkara et al. and Steward et al. [41, 40].

This holds true for every patient, even though some will have a reduced ovarian reserve, and will thus not be able to arrive at this range. For these patients, the dose will be adjusted as needed to bring them as close as possible to the optimal range. To note: we have described dose as *d*_*i*_∈ [0, ∞), hence there is a minimum set by definition. But this minimum can be higher than 0, and it is very likely that a maximum limitation exists. Therefore, it is not only physiologically difficult for some patients to get to the optimal outcome range, patients can also be limited by the range of available doses depending on the use case. In this specific use case, *d*_*min*_ has been set at 100 IU of FSH, and *d*_*max*_ ranging from 300 to 450 IU has been explored.

### 5.1. IDoser for FSH dosing

There are two elements essential for the application of our proposed IDoser in all cases: the core model and the loss function. For this study, the core dosing model selected assumes an underlying linear dose-response and is defined as

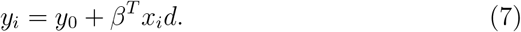

Then, given a desired outcome *y*^*\**^, it can be rearranged into our dosing model 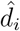 as follows

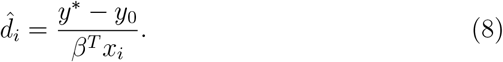

This can be generalized to the following dosing model

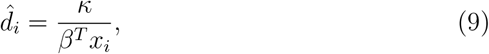

which will have as parameter set *γ* both *κ* and *β*, that is *γ* = (*κ, β*).

For the loss function, additional rules (outside the basic ones described) were defined to ensure an improved but conservative dosing policy, as highly variable doses are discouraged due to greater uncertainty regarding the expected outcome. Limitations in dose changes were defined depending on the outcome range for the specific patient. Following the definitions by Polyzos and Sunkara [34], the next categories where defined:

- An outcome below than 4 mature oocytes was considered too low;
- An outcome between 4 and 9 mature oocytes was considered suboptimal;
- An outcome between 10 and 15 mature oocytes was considered optimal; and
- An outcome greater than 15 mature oocytes was considered too high.

Accordingly, higher changes were allowed for patients with a too low and too high outcome compared to those with a sub-optimal outcome. Specifically, a dose modification up to 150 IU was allowed in the first two instances, and up to 75 IU for those in the latter cases. Changes up to 25 IU were not considered as such. All these thresholds were established in collaboration with expert professionals in the field.

## 6. Evaluation Methodology

### 6.1. Literature benchmark

We identified from the existing literature an implementation described in the study by La Marca et al. [26] and later tested via an RCT [2]. This work uses a core model similar to the one in our research, ensuring positive monotonicity. Additionally, our second assumption was referenced in the paper by fixing *y*^*\**^ to 9 oocytes for all patients, leaving implicitly our concept *y*_0_ equal to 0. Thus, we decided to use it as the literature *benchmark* for our study, and from here onward will be referred to as La Marca or LM.

Their covariates included age, AMH and FSH. The developed and published model was derived from running a linear regression of the following equation:

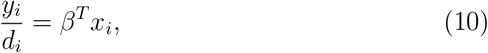

where the coefficients included in *β* were estimated to construct the dosing model. The dosing model constructed then would be expressed as

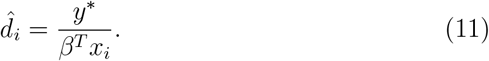

In the following RCT [2] a significantly higher proportion of patients got an optimal outcome (described here as 8 to 14 oocytes), even if the mean number of oocytes was not significantly changed.

### 6.2. Optimization exploration

Several approaches were explored when optimizing the LM model, described in Appendix A, and compared statistically to clinical practice and the unmodified LM model in Appendix B. The final model was obtained after including two extra covariates available in our dataset (AFC and BMI), and omitting the variable basal FSH. All parameters of *γ* were optimized and used for the final proposed doser.

### 6.3. Model comparison and statistic tests

To compare the optimized models to LM and clinical practice two methods were used. The first one was analyzing and plotting 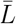 across all the *d*_*max*_ values allowed. In this first method, an extra comparison was performed to understand the quality of the IDoser and LM models. This extra comparison introduced an oracle decision policy or model, where the doses recommended are always correct: if a dose change is needed it is done in the right direction and inside the adequate range. This is done by simply determining if the outcome *y*_*i*_ is inside or outside the optimal range. If it is outside, a dose change is needed, and it is executed in the correct direction and range. Hence, the oracle model’s dose recommendations represent all available and correct dose changes for the test patients, or in other words, a perfect policy as per our loss function. The 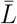 value for the oracle indicates the maximum improvement possible in the validation dataset.

To test if any of the methods were statistically different from clinical practice or among themselves, loss values from every group were compared using the method recommended in García and Herrera [13] and García et al. [12], which are an extension of the study by Demsar [10]. Specifically, Iman-Davenport’s corrected Friedmann test [18] (as the normality test failed) was used. When significant results were achieved, a post-hoc test was used to determine which groups were different. P-values were adjusted using Finner’s correction, as per García et al. [12]. R’s package *scmamp* was used to run the mentioned tests. A p-value of less than 0.05 was considered significant.

## 7. Results

As aforementioned, the first methodology used to compare our proposed model, IDoser, to LM and clinical practice was to plot 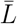 of every model after they were used to dose our validation dataset across all 4 *d*_*max*_ explored. Clinical practice was always described always as *L* = 0. The values obtained were plotted in Figure 4, together with the oracle’s value (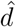 always in right direction and range).

**Figure 4:**
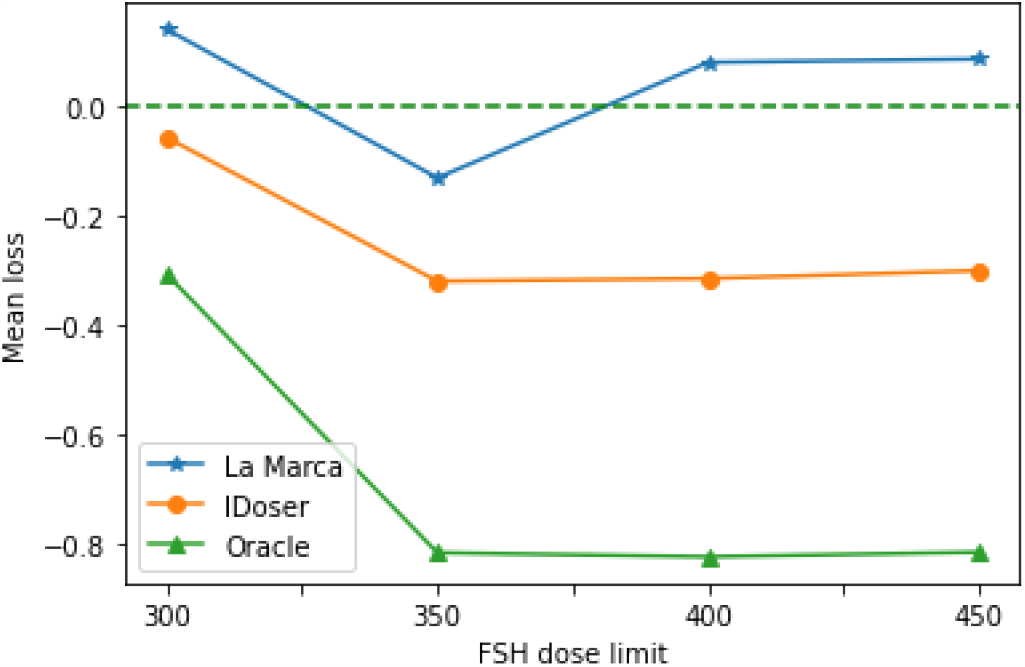
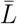 across *d*_*max*_ for La Marca, oracle and the proposed IDoser when used to dose in the validation dataset. The dashed line marks *L* for the clinical practice dosing method.

As it can be clearly observed, the IDoser is always under LM 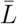 values and below the 0 mark, where clinical practice lies. It is also clear that the oracle model lies far below both of them, indicating that there is still a gap to be filled.

Regarding statistical results, Iman-Davenport’s corrected Friedman test results (shown in Table 2) prove a significant difference between models’ loss across all selected points of *d*_*max*_.

**Table 2:** Results of Iman Davenport’s correction of Friedman’s rank sum test of all methods tested across the 4 selected values for *d*_*max*_

| $d_{max}$ | 300 | 350 | 400 | 450 |
| --- | --- | --- | --- | --- |
| **p-value** | 0.002657* | 4.977e-11* | 5.373e-14* | 8.36e-14* |

The consequent post-hoc test results showed a significant improvement of our optimized model compared to the LM model across all *d*_*max*_ points explored. This also holds true when compared to clinical practice, except in the case of *d*_*max*_ = 300, where even if an improvement is observed, it cannot be proven statistically. These results are represented in Table 3. Specific 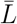 and adjusted p-values are listed in Tables 4 to 7 in Appendix A.

**Table 3:** Ordered results from worst (left) to best (right) method in one vs one comparison across all *d*_*max*_ values. Results extracted from post-hoc test with p-values adjusted by Finner’s methodology.

| $d_{max}$ | Ordered results by significant differences |
| --- | --- |
| **300** | La Marca $\prec$ Clinical Practice ∼ IDoser |
| **350** | Clinical Practice $\prec$ La Marca $\prec$ IDoser |
| **400**<br>**450** | La Marca ∼ Clinical Practice $\prec$ IDoser |

**Table 4:**
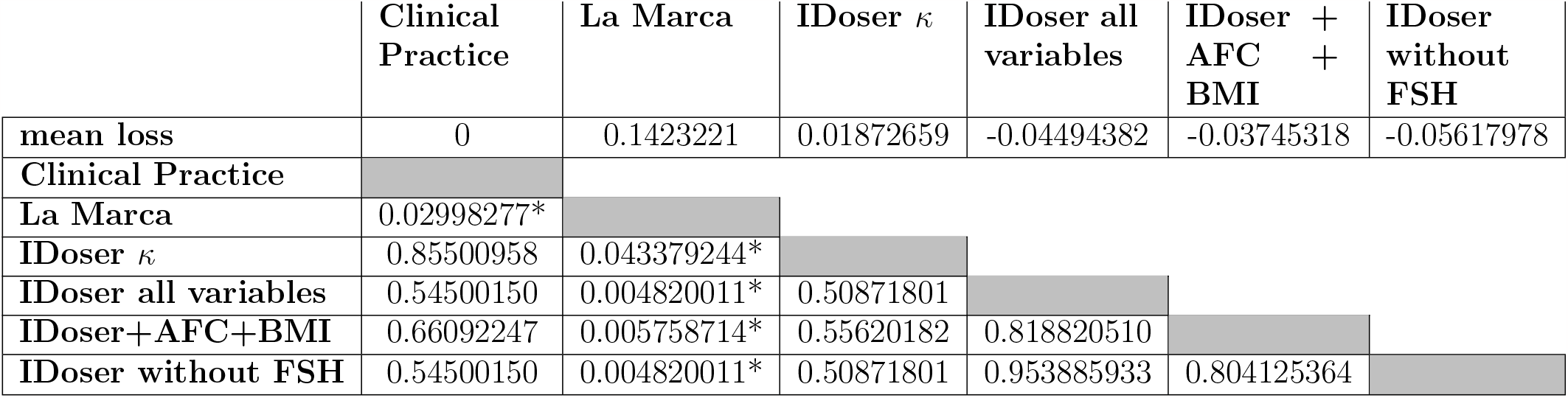
Posthoc test of differences in individual losses between explored methods and clinical baseline capping *d*_*max*_ at 300, p-values adjusted using Finner method and marked with * when under 0.05.

**Table 5:**
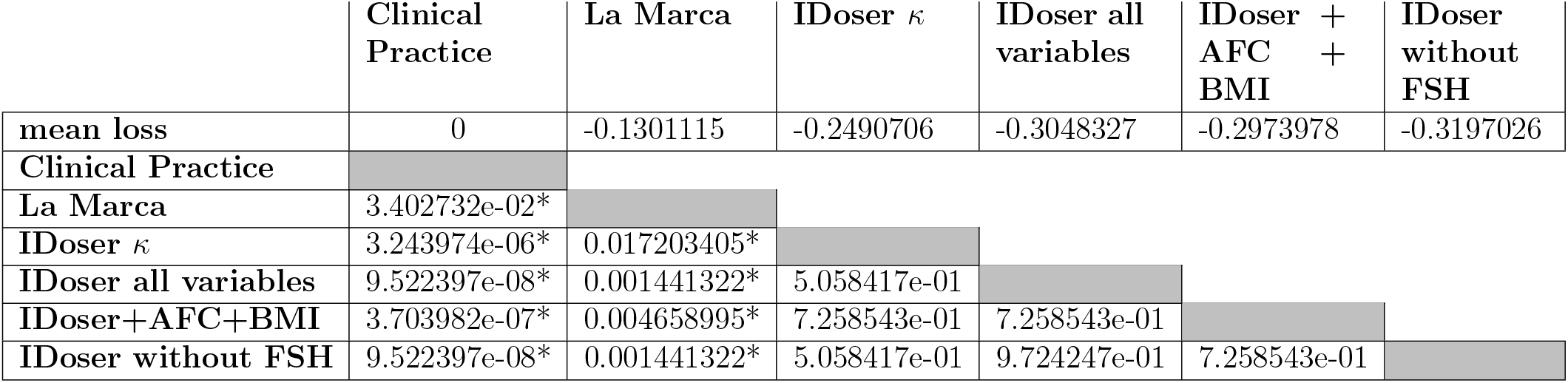
Posthoc test of differences in individual losses between explored methods and clinical baseline capping *d*_*max*_ at 350, p-values adjusted using Finner method and marked with * when under 0.05.

**Table 6:**
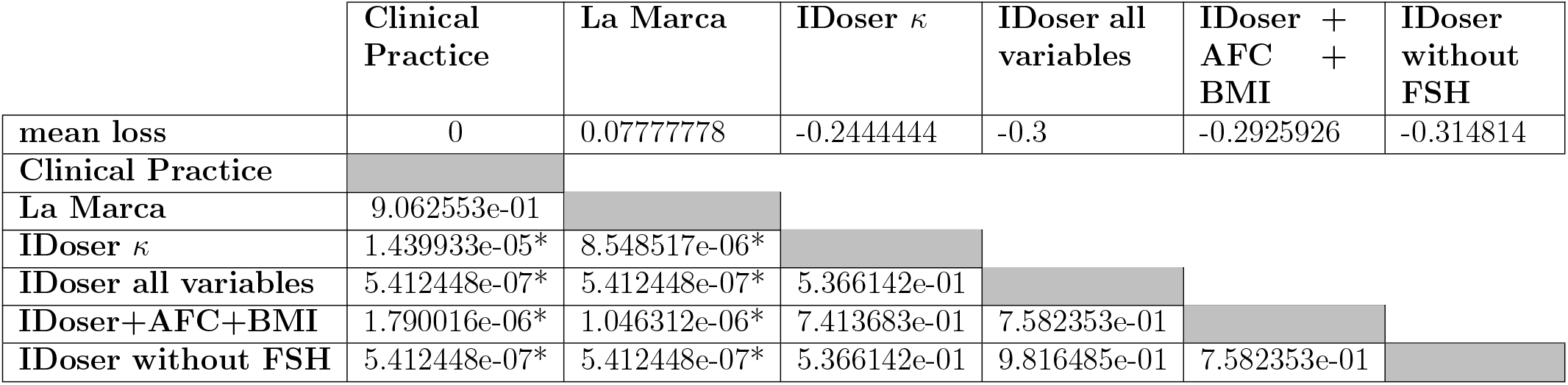
Posthoc test of differences in individual losses between explored methods and clinical baseline capping *d*_*max*_ at 400, p-values adjusted using Finner method and marked with * when under 0.05.

**Table 7:**
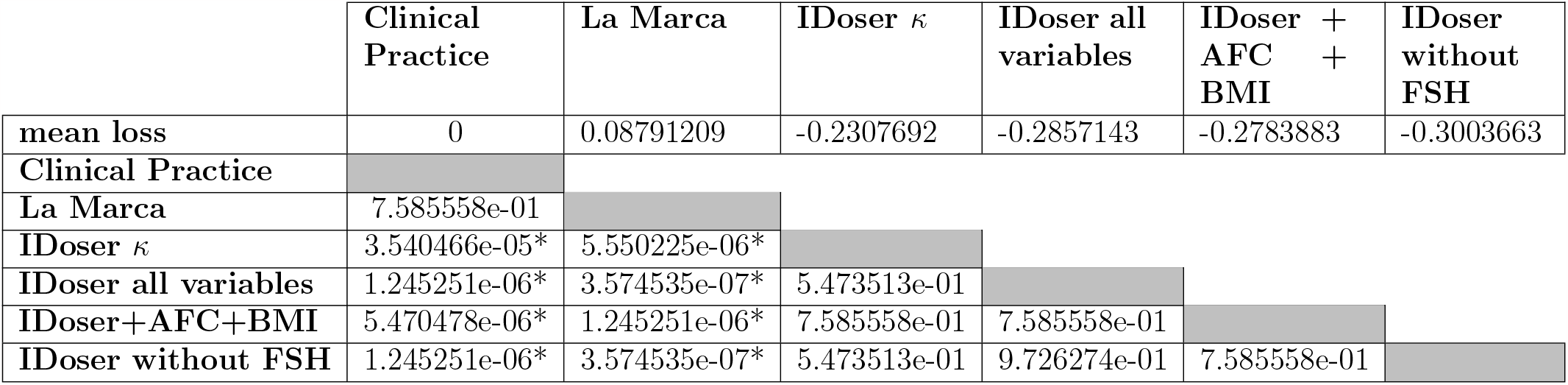
Posthoc test of differences in individual losses between explored methods and clinical baseline capping *d*_*max*_ at 450, p-values adjusted using Finner method and marked with * when under 0.05.

## 8. Discussion

The proposed IDoser model achieved a significant improvement compared to the LM model across all investigated *d*_*max*_ values, and most of the times also when compared to baseline clinical practice or policy, except for *d*_*max*_ = 300. Here, the improvement achieved did not reach the significance threshold stipulated. As shown on Figure 4, there is actually less margin for improvement in dosing policy compared to the rest of values of *d*_*max*_. This would explain why, with our current sample size, a significant difference compared to clinical practice with *d*_*max*_ = 300 can not be shown, as there are few cases that can be improved, and IDoser does not identify a correct dose change for all of them. This clearly evident from the distribution of improvable cases across the outcome axis (number of mature oocytes), most of which are concentrated in low or sub-optimal ranges (Figure 5).

**Figure 5:**
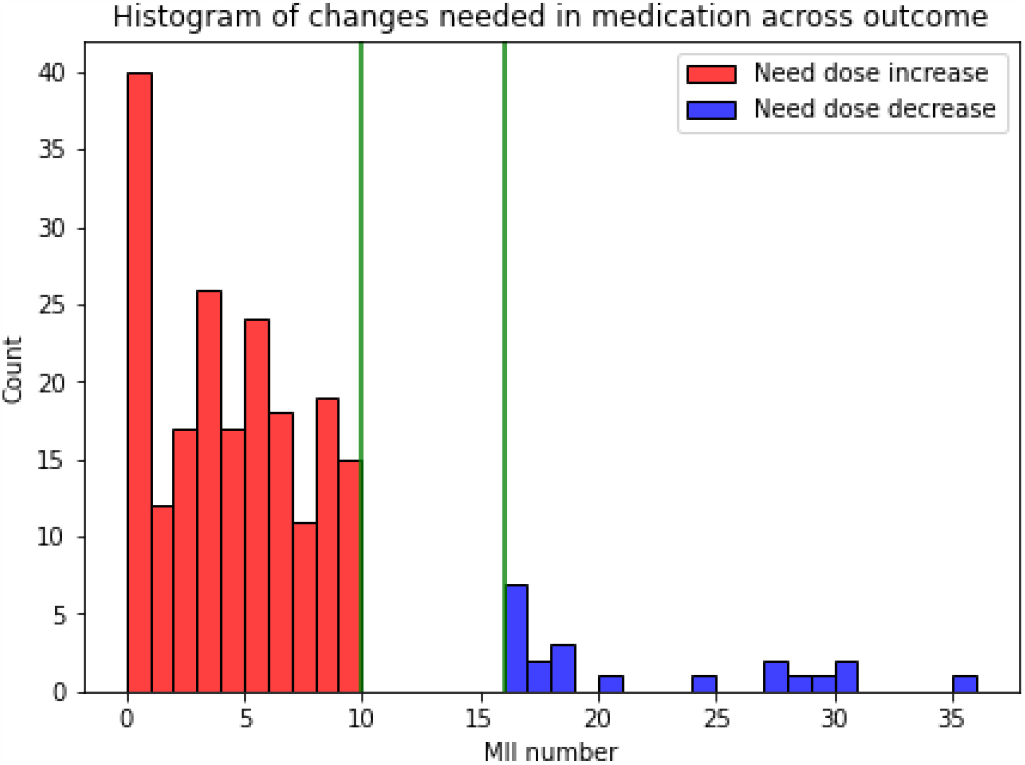
Distribution of cases that need an increase of dose (red) or decrease (blue) for the validation dataset if *d*_*max*_ = 450 is allowed.

These cases would need a significant increase of medication, however, as expected, many low-responder patients have already received 300 IU of FSH by their clinician. This value of *d*_*max*_ is most commonly used, supported by the new European Society of Human Reproduction and Embryology (ESHRE) guidelines for ovarian stimulation [42]. As shown in Figure 6, there are still some cases in the validation database that have been dosed over 300 IU in clinical practice, indicating that clinicians thought that some specific patients may benefit from exceeding the broadly recommended *d*_*max*_, as studies used for the guideline are based on population tendencies and not individuals. Given that the IDoser model has been optimized to also auto bound itself for a more conservative dosing policy (with an optimized *d*_*max*_ of 333 IU), it could be used safely with an open *d*_*max*_ in order to identify which patients are estimated to be candidates for a FSH dose over 300 IU of FSH. Figure 6 also aids to visualize how IDoser smooths and shifts the dose distribution versus clinical practice slightly upwards, and how it is limited to 333 IU by its automatic bounding system. On the contrary, the LM model tends to distribute doses more evenly (not as centered in 300), having more cases with decreased doses and doses over 300 concentrated in the 450 IU mark (*d*_*max*_ value implemented in Figure 6).

**Figure 6:**
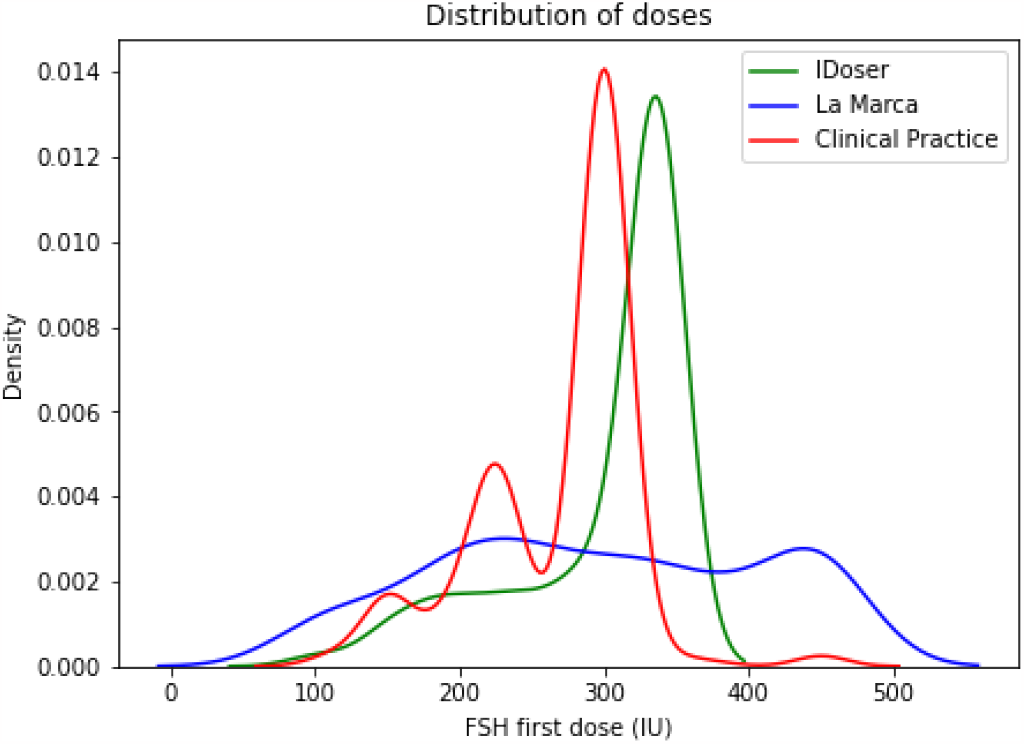
Distribution of doses for La Marca (blue), Clinical Practice (red) and IDoser (green) for the validation dataset with *d*_*max*_ = 450

These antagonistic tendencies can also be clearly visualized in Figures 7 to 10, where dose changes distributed across outcome are shown for both models (IDoser and LM) and *d*_*max*_ 300 and 450. IDoser (Figures 7 and 9) tends to rescue more cases under our defined 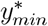, where the majority of improvable cases lie, at the expense of very few patients over 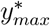 having their dose increased and some not decreased. This could be due to an under-representation of this subset of patients in our dataset, and should be considered a limitation of the resulting model. On the contrary, the LM model performance (Figures 8 and 10) shows that more patients that need a reduction on dose get it, but at the same time many patients that need an increase in dose are instead given a reduced one. That is clearly why our loss function is penalizing this model.

**Figure 7:**
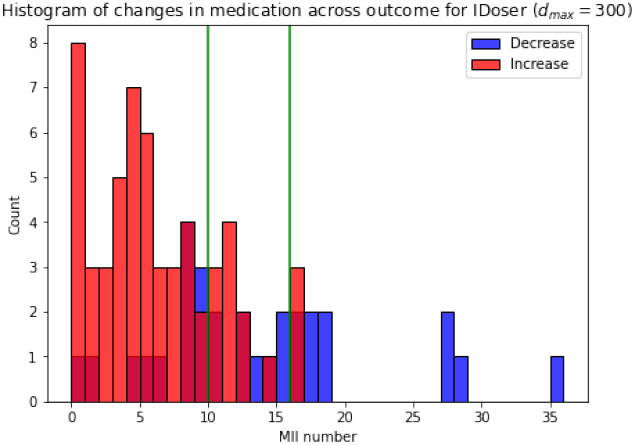
Doses changes for the IDoser model with *d*_*max*_ = 300

**Figure 8:**
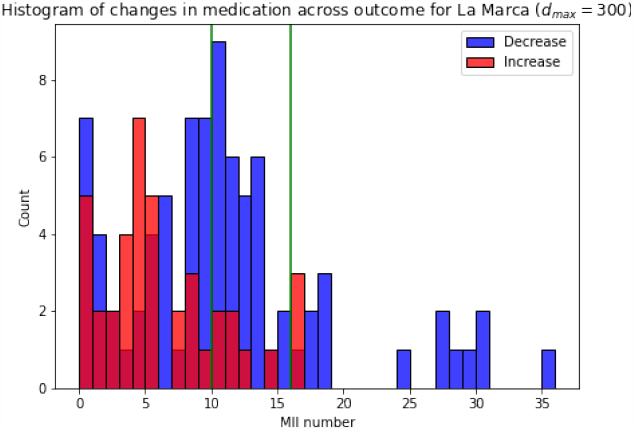
Doses changes for the La Marca model with *d*_*max*_ = 300

**Figure 9:**
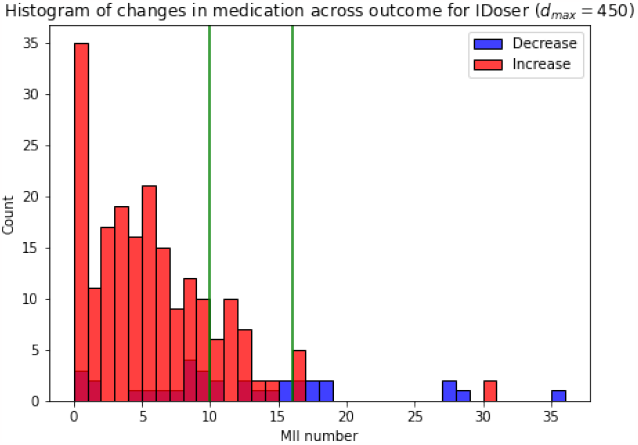
Doses changes for the IDoser model with *d*_*max*_ = 450

**Figure 10:**
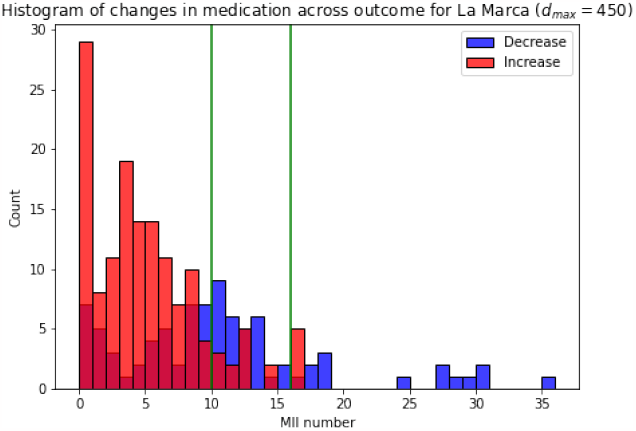
Doses changes for the La Marca model with *d*_*max*_ = 450

It could be argued that the decreasing tendency of the LM model could be derived by its use of a *y*^*\**^ value of 9, one step lower than our defined 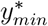 of 10. Of course, this could be the case for some patients in our databases, but as shown in Figures 8 and 10, it would not explain the relevant dose reductions in patients with very low outcomes. Another cause of the lower performance of the LM model could be that it was originally developed for normo-ovulatory patients under 40 years, and our database comprises all patients eligible for an IVF treatment. That characteristic was the motivation for this study, as all published FSH dosing models excluded critical portions of the patient population. Also notable, is the vantage obtained with IDoser while not using a single point for *y*^*\**^, but a range of desired outcomes. This makes a model less prone to change doses in the desired interval, as shown in Figures 7 to 10.

Finally, it is also worth noting that our loss function penalizes dose changes considered to be too big, even though they may be in the right direction. Given that our model has been optimized with these rules to avoid a model that is too bold, the fact that it still recommends significant changes in dose could be due to some patients truly needing such a change. Importantly, true clinical utility of IDoser can only be proven by a prospective randomized trial.

The methodology described here may be applied to any similar dosing problem. Existing methodologies, such as causal inference adn double machine learning are very interesting and robust, but in cases like the one described in this study, are far from applicable. Nevertheless, there is still a need to improve dosing policies using evidence-based knowledge for the sake of patients that receive subpar clinical care following generalized policies.

Observational datasets available in clinical practice, generated using dosing protocols, are prone to have little variability. Moreover, not all confounders can be accounted for evenly across all cases, as different clinicians favor different biomarkers or prescribe more or less complete test batteries depending on experience or other factors, including financial considerations or patient request. Hence, algorithms trained on those databases may not conform with evidence-based knowledge, and even sometimes plain logic. It is clear that formalizing the rules that a clinician applies based on their experience, and packaging them in a selected core model and a loss function can help in the process of obtaining a model that follows those rules. These two rather simple elements allow for significant versatility for implementation in different dosing settings (negative monotonicity, changing core function, etc.). These concepts partially take inspiration from PK/PD modelling, where physiological and pharmacological assumptions and principles are followed. This is translated into our methodology by including a core model were the monotonic assumption is heeded, and by including rules in our loss function that penalize any dose change in the wrong direction. In this study, we explore a core function derived from a linear dose-response. Other functions, including exponential and a straight linear function relating dose to covariates were investigated, however the one presented gave the best results. Other core models that are closer to physiological dose-response relationships, such as sigmoid functions may be explored in the future.

As IDoser is only applicable to single-dose dosing cases, and does not take into account adjustment of dose over time for each individual patients.

Ultimately, IDoser achieved an optimized dosing policy in a time-efficient manner, but can also be implemented in a conservative way to validate drug doses “in silico”. This is especially important, given that RCTs entail a significant investments both in time and money. Being able to demonstrate some expected improvement non-interventionally ensures a faster acceptance of the route of an RCT.

IDoser constitutes a clear and straightforward method to implement evidencebased knowledge to train individualized dosing models with a relevant predicted improvement on current clinical practices. This is especially relevant, as in several instances the historical databases available are not amenable to more complex methodologies.

Future lines of work include the implementation of individual values for *y*^*\**^, the use of different optimization methodologies outside of coordinated descent, or more complex core models close to real dose-response functions.

## Data Availability

All data produced in the present study are available upon reasonable request to the authors

## 9. Acknowledgments

The authors declare no conflicts of interests. Permission to conduct this study was obtained from the Ethical Committee for Research of Eugin on 20 October 2020 (approval code: ALGO2).

This work was supported by Doctorat Industrial funded by Generalitat de Catalunya [DI-2019-24], by project CI-SUSTAIN funded by the Spanish Ministry of Science and Innovation [PID2019-104156GB-I00], by EUROVA Innovative Training Network (MSCA- ITN-2019-860960), and by intramural funding by Clínica Eugin-Eugin Group.

Nuria Correa is a PhD Student of the doctoral program in Computer Science at the Universitat Autonoma de Barcelona.

We would like to thank Dr Daniel Mataró Marçal for kindly lending us his time and his expertise on ovarian stimulation protocols.

## 10. Author contributions

All authors have contributed in the different aspects of this study. Nuria Correa leaded the conceptualization, data curation, experimentation, and writing of the paper as part of her PhD.

## Appendices

### A. Optimization exploration

Given that by our definition 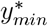 and 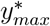 are slightly higher (10 to 15) we primarily used their *β* coefficients and optimized only *κ*. Next, our workflow was designed to explore optimization of all coefficients in *γ* and addition/omission of covariates. Specifically:

1. Optimization of only *κ*
2. Optimization of all *γ*
3. Addition of AFC and BMI covariates, available in our database
4. Omission of basal FSH covariate

This resulted in 4 new optimized dosing models to be compared to the benchmark and the clinical dosing policy recorded in our database, which we will refer to from now on as *baseline*. Once *γ* values were obtained for all 4 models, a secondary optimization was run to automatically find an upper bound to dose or 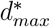, as a further measure for a safe and conservative model. Every model was trained with all available data depending on the covariates included, but validated always on the same database where all covariates were filled in, to avoid possible biases on the population. The benchmark and our 4 iterations were validated across 4 possible *d*_*max*_: 300, 350, 400, and 450. For each limit, validation cases with *d* up to that value were admitted, and every model was allowed to dose at a maximum of the same value, and every individual loss evaluated. It is worth noting here that our models were autolimiting themselves with their optimized value of 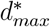, whenever this value was lower than any of 4 *d*_*max*_ explored.

In the end, only the best of all 4 iterations was selected as our final proposal.

### B. Statistics results

## Notes

### Competing Interest Statement

The authors have declared no competing interest.

### Author Declarations

Ethics Committee for Research of Clinica Eugin gave ethical approval of this work (approval code: ALGO2) on the 20th of October of 2020.

